# Cohort Profile Update: GendAge and Berlin Aging Study II (BASE-II)

**DOI:** 10.1101/2020.09.05.20187898

**Authors:** Ilja Demuth, Verena L. Banszerus, Johanna Drewelies, Sandra Düzel, Ute Seeland, Dominik Spira, Esther Tse, Julian Braun, Lars Bertram, Andreas Thiel, Ulman Lindenberger, Vera Regitz-Zagrosek, Denis Gerstorf, Additional BASE-II/GendAge investigators

## Abstract

The study “Sex- and gender-sensitive prevention of cardiovascular and metabolic disease in older adults in Germany”, the GendAge study, focusses on major risk factors for cardiovascular and metabolic diseases and on the development of major outcomes from intermediate phenotypes in the context of biological sex and gender differences. It is based on a re-investigation of participants of the Berlin Aging Study II (BASE-II). The Berlin Aging Study II (BASE-II) is aiming at identifying factors that distinguish healthy from unhealthy ageing and completed baseline assessments in 2,200 adult volunteers (1,600 participants aged 60–80 years and 600 participants aged 20–35 years) in 2014. The BASE-II follow-up assessments of 1,100 men and women aged 65–94 years in 2018–2020 were part of GendAge. In addition to re-assessing most baseline measures (geriatrics, internal medicine, immunology and psychology) we implemented a comprehensive gender questionnaire covering socio-cultural gender characteristics and added high-quality echocardiography.

## The original BASE-II cohort

The Berlin Aging Study II (BASE-II) was launched as a multidisciplinary study aiming at identifying factors that distinguish ‘healthy’ from ‘unhealthy’ ageing. Baseline recruitment of 2,200 adult volunteers from the Berlin metropolitan area and baseline assessments were completed in 2014^1^. The ascertainment protocol included the collection of data from different domains for each of the 1,600 participants aged 60–80 years and 600 participants aged 20–35 years, namely geriatrics and internal medicine, immunology, genetics, psychology, sociology, and economics^1,2^.

BASE-II baseline data were used in a multitude of analysis projects focusing on key questions revolving around age and aging. Research topics of the ongoing study include, but are not limited to, cognitive aging^3–5^, cardiovascular and metabolic health^6–8^, sarcopenia and frailty^9,10^, psychosocial factors of aging^11,12^, genetic risk factors of aging and disease^13–15^, the impact of characteristics of the neighborhood people are living in^16^, as well as indicators of biological age^17,18^ and immune biomarkers^19^. For an overview of the BASE-II research foci and publications, refer to ^20^ and the BASE-II website (https://www.base2.mpg.de/en/project-information/publications).

## What is the reason for the new data collection?

The study “Sex- and gender-sensitive prevention of cardiovascular and metabolic disease in older adults in Germany”, the GendAge study, focusses on major risk factors for cardiovascular and metabolic diseases and on the development of major outcomes from intermediate phenotypes in the context of biological sex and gender differences. Major outcomes include but are not limited to myocardial infarction (MI), heart failure (HF) and T2D, as well as mortality and quality of life. Major aim is the first systematic collection of follow-up data in BASE-II participants and the analysis of sex- and gender-related differences. Gender was obtained by a novel comprehensive gender questionnaire covering a range of socio-cultural gender characteristics as a central instrument. This questionnaire contains an adapted version of the gender questionnaire developed by Pelletier and colleagues^21^ and additionally included the variables used to calculate a gender score retrospectively by making use of gender-related variables assessed at baseline^22^. This gender questionnaire will be central to develop a second gender score (GS-II).^23^.

## What will be the new areas of research?

There is new knowledge showing that sex differences play a role in all major diseases, their prevention and treatment^24^. Other studies showed that gender as the socio-cultural dimension of disease affects disease and treatment outcomes and also well-being^21,25^. The new areas of research cover the systemic inclusion of sex-specific analysis and the inclusion of gender. Aging interacts with sex and gender differences in health, but it is not clear, which mechanisms are most important.

GendAge aims to better understand, which mechanisms affect cardio-metabolic morbidity, mortality, and quality of life among older adults in a sex- and gender-sensitive manner. While on different occasions follow-up data were ascertained for questionnaire and cognitive data^5,26–30^, as being part of the GendAge study this cohort profile update describes, the first comprehensive follow-up assessments in BASE-II that also includes a re-assessment of central variables in the areas of internal medicine and geriatrics. At the same time, the other BASE-II research foci established over the past 10 years as described above (and in^20^) will be continued and strengthened by the transition of BASE-II into a longitudinal study.

## Who is in the cohort?

The original BASE-II sample consisted of 2,200 participants from the greater metropolitan area of Berlin, Germany (baseline assessment between 2009 and 2014). The follow-up assessments within the GendAge study took place between 22 June 2018 and 10 March 2020 at the Charité Universitaetsmedizin Berlin. During the recruitment of the follow-up cohort, we approached all participants of the remaining pool of 1,428 subjects out of the originally 1,671 subjects who completed the baseline medical assessments at an age of 60 years and older. Potential follow-up participants were contacted via telephone and an invitation letter containing the comprehensive participant’s information sheet and the letter of consent was sent at least five days before the scheduled first of two assessment days to all subjects who agreed to participate at least five days before the scheduled first of two assessment days. Additionally, we included 17 subjects of the older age group who were not medically assessed at baseline, but were part of the BASE-II sample with data collected from at least one of the BASE-II partner sites. As presented in the flow chart (Figure 1), this resulted in a total of 1,100 participants of the older BASE-II group investigated in the followup. These older participants had a mean age of 75.60 years (SD ± 3.77, range 64.91 – 94.07 years), with up to 10.37 years of follow-up (N = 1,083, mean follow-up at 7.35 years, (SD ± 1.46), range 3.91 – 10.37). At baseline, the BASE-II included a group of 600 younger subjects aged 20–35 years serving as a reference population^1^, of which 500 completed baseline medical assessments. Between 7 February 2020 and 13 March 2020, we performed followup assessments in a total of 64 participants of this younger group until these assessments were suspended because of the SARS-Cov2 pandemic. These younger participants had a mean age of 36.81 years (SD ± 3.46, range 29.32 – 44.05 years), with up to 10.7 years of follow-up (minimum 6.08 years, mean follow-up at 8.16 years, SD ± 1.58).

**Figure 1:**
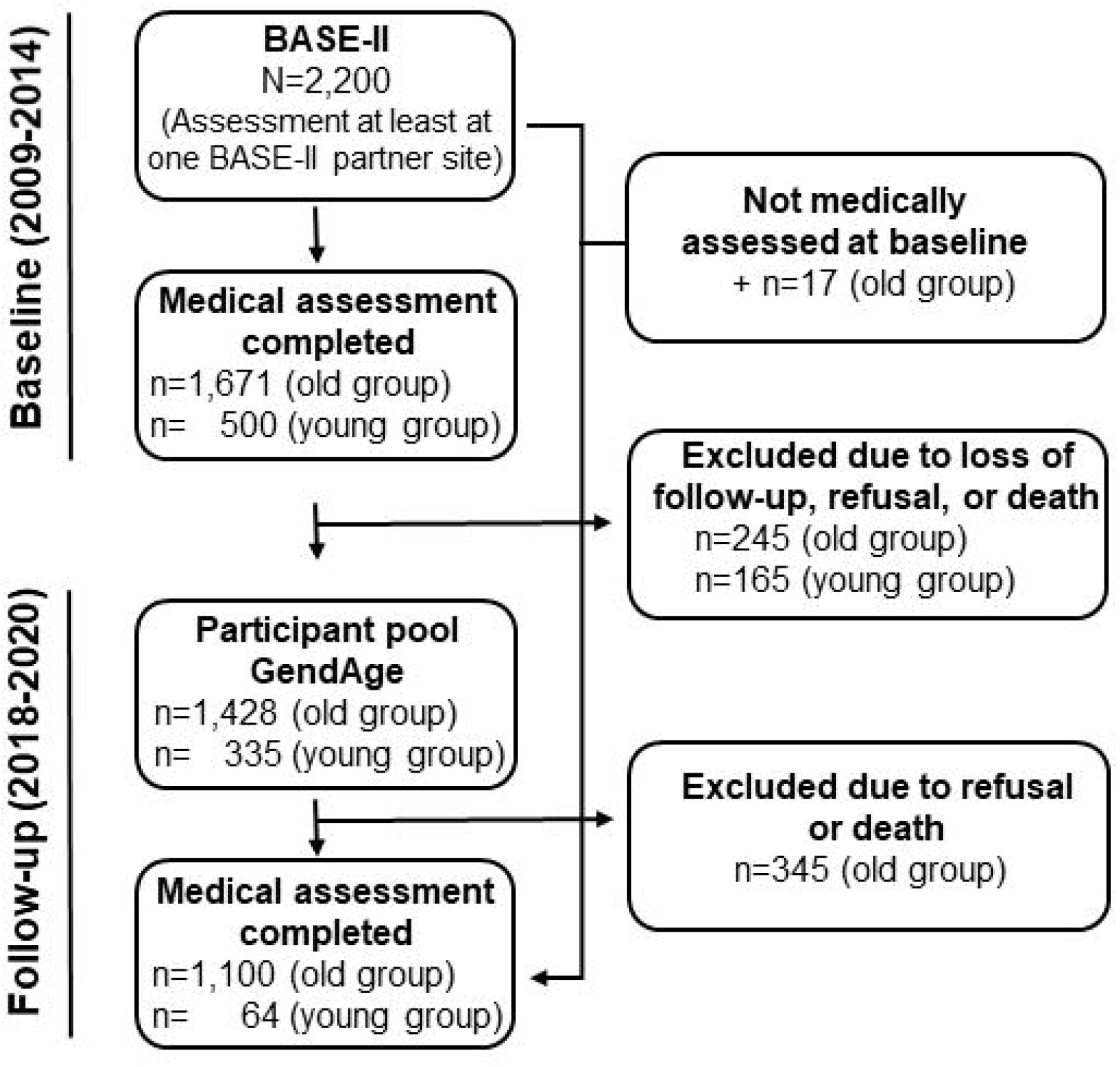
Flow chart explaining the final BASE-II sample with follow-up assessments completed in GendAge

At follow-up, almost all of the older participants were retired (97.3%) as compared with 86% at the time of baseline assessment. At baseline, BASE-II participants were characterized by higher education and better self-reported health status than the general population of Berlin and Germany^1^. At follow-up, this selection seems to have increased, with 68.8% of the participants reported to have a high school degree (51% at baseline) and about 61% rated their health as *very good* or *good* (40% at baseline). The rate of divorce had been above average at baseline with 29% and had dropped to 21.7% at follow-up, which is still significantly above the German and Berlin average (i.e. 12.0% and 17.4%, respectively)^31^, while the proportion of widowed participants increased from 5% at baseline to 10.5% in the follow-up dataset of older BASE-II participants. As shown in Table 1, differences between men and women are evident with respect to the sociodemographic status and psychosocial functioning in the follow-up cohort: Men reported significantly higher school degrees and higher satisfaction with life in general than women. Interestingly, self-rated health did not differ between men and women, which matches to the overall morbidity estimated by an adapted version of the Charlson morbidity index^17,32^, which also did not differ between men and woman (p = 0.981, Table 1). This morbidity index, however, increased between baseline and follow-up (p< 0.001, Wilcoxon signed-rank test and data not shown). Differences between men and women exist in the follow-up dataset with respect to the prevalence of some, but not all cardiovascular risk factors and diseases (Table 1). Men for example had a higher BMI and a higher proportion of men reported to have T2D and myocardial infarction than women. No significant differences between men and women were evident in the reporting of hypertension, peripheral artery disease, and stroke. With the aim of achieving a particularly high quality standard in the assessment of participant’s medical history at baseline and follow-up, including past and current diseases, the information given by the participants was recorded from study physicians as part of a structured one-to-one interview, allowing to consider its plausibility. This, however, does not cover the gap between reported (anamnestic) diseases and the diseases diagnosed in the course of the study. This is exemplified by T2D, which was reported by 15.6% and 8.8% of men and women, respectively. In contrast, this disease was diagnosed in 20.7% of men and 13.3% of women based on the ESC criteria 2019^33^, indicating that a substantial proportion of almost 30% was unaware of the disease. The T2D prevalence increased from 12.7% at baseline to 16.7% at follow-up in the total cohort.

**Table 1:** Selection of BASE-II follow-up characteristics as assessed of the GendAge study – older group of participants

|  | **Total number of observations** | **Women<sup>1</sup>**<br>**(N=573, 52.1%)** | **Men<sup>1</sup>**<br>**(N=527, 47.9%)** | **p-value<sup>2</sup>** |
| --- | --- | --- | --- | --- |
| **Age (years)** | 1,100 | 75.7 (± 3.5) | 75.5 (± 4.0) | 0.276 |
| **Highest school degree**<br>Elementary school<br>Intermediate school<br>High school | 1,095 | 35 (6.1%)<br>183 (32.0%)<br>354 (61.9%) | 18 (3.4%)<br>104 (19.9%)<br>401 (76.7%) | 8.4×10<sup>-7</sup> |
| **Family status**<br>Married<br>Not married, in partnership<br>Single<br>Divorced<br>Widowed<br>Other | 1,098 | 218 (38.0%)<br>12 (2.1%)<br>60 (10.5%)<br>187 (32.6%)<br>89 (15.5%)<br>7 (1.2%) | 386 (73.5%)<br>19 (3.6%)<br>33 (6.3%)<br>51 (9.7%)<br>26 (5.0%)<br>10 (1.9%) | 3.0×10<sup>-34</sup> |
| **Employment status**<br>Retired | 1,055 | 540 (97.6) | 486 (96.8) | 0.689 |
| **Self-rated health**<br>Very good<br>Good<br>Fair<br>Poor or very poor | 1,096 | 56 (9.8%)<br>284 (49.7%)<br>166 (29.0%)<br>66 (11.5%) | 65 (12.4%)<br>262 (50.0%)<br>143 (27.3%)<br>54 (10.3%) | 0.499 |
| **Satisfaction with life in general** | 1,097 | 7.9 (±1.6) | 8.1 (±1.4) | 0.034 |
| **Digit Symbol Substitution Test<sup>3</sup>** | 1,095 | 41.82(±8.28) | 39.65(±8.95) | 3.4×10<sup>-5</sup> |
| **Depression (ever diagnosed)** | 1,095 | 122 (21.3%) | 63 (12.0%) | 5×10<sup>-5</sup> |
| **BMI** | 1,098 | 26.6 (±4.7) | 27.4 (±3.7) | 0.005 |
| **Diabetes mellitus type II (self-reported)** | 1,097 | 49 (8.6%) | 82 (15.6%) | 3.7×10<sup>-4</sup> |
| **Diabetes mellitus type II (diagnosed/ ESC criteria 2019)** | 1,097 | 76 (13.3%) | 109 (20.7%) | 0.001 |
| **Metabolic Syndrome (diagnosed, AHA/IDF/NHLBI criteria 2009)** | 1,074 | 252 (45.5%) | 327 (62.9%) | 1.2×10<sup>-8</sup> |
| **Hypertension** | 1,097 | 296 (51.7%) | 311 (59.2%) | 0.013 |
| **Myocardial infarction** | 1,097 | 11 (1.9%) | 24 (4.6%) | 0.015 |
| **Stroke** | 1,096 | 13 (2.3%) | 20 (3.8%) | 0.158 |
| **Peripheral artery disease** | 1,094 | 8 (1.4%) | 15 (2.9%) | 0.138 |
| **Morbidity index** | 955 | 1.0 (IQR 2.0) | 1.0 (IQR 2.0) | 0.594 |
| **Pulse wave velocity (m/s)** | 932 | 11.21 (±0.92) | 11.04 (±0.91) | 0.006 |
| **Left ventricular ejection fraction (%)** | 774 | 64.16 (±6.42) | 62.90 (±5.75) | 0.004 |
| **Frailty (Fried)**<br>Not frail<br>Pre-frail<br>Frail | 1,087 | 260 (45.4%)<br>280 (48.9%)<br>28 (4.9%) | 251 (47.6%)<br>248 (47.1%)<br>20 (3.8%) | 0.542 |
| **Maximal hand grip strength (kg)** | 1,098 | 20.5 (±4.4) | 35.1 (±6.8) | $3.1 \times 10^{-236}$ |
| **Recent thymic emigrants (naïve CD4<sup>+</sup> T cells)** | 395<sup>3</sup> | 64.69 (± 16.34) | 51.03 (± 13.69) | $6.3 \times 10^{-16}$ |
| **T<sub>EMRA</sub> (effector memory T cells re-expressing CD45RA)** | 395<sup>4</sup> | 32.82 (± 19.96) | 34.94 (± 21.29) | 0.309 |
| **Cytotoxic SLAMF7+ CD4<sup>+</sup> T cells** | 181<sup>5</sup> | 6.02 (± 5.98) | 6.05 (± 6.60) | 0.974 |

<sup>1</sup>Data are presented as N (%), mean ± standard deviation or median (interquartile range, IQR). <sup>2</sup>Differences between women and men were assessed using the parametric t test, the nonparametric Mann-Whitney U test or the $\chi^2$ where appropriate. <sup>3</sup>assessed at study visit 1 <sup>4</sup>903 expected to be available after completion of the analyses. <sup>5</sup>700 expected to be available after completion of the analyses.

## What has been measured?

With the aim of investigating human ageing processes in BASE-II under consideration of different disciplines and longitudinally, the baseline investigation aimed at the most comprehensive data collection possible. At follow-up, most of these data in the field of geriatrics, internal medicine and psychology were again part of the study protocol (for a select overview, see Table 2). As the follow-up assessment being part of the GendAge study, we have implemented a comprehensive gender questionnaire covering a range of sociocultural *gender* characteristics as a central instrument. This questionnaire contains an adapted version of the gender questionnaire developed by Pelletier and colleagues^21^ and additionally included the variables used to calculate a gender score retrospectively by making use of gender-related variables assessed at baseline^22^. This gender questionnaire will be central to develop a second gender score (GS-II) and to reach the GendAge goals.

**Table 2:** BASE-II follow-up assessments during the three study visits in the GendAge study

| Type of assessment/<br>domain | Example assessments/tests |
| --- | --- |
| Physical examination<br>and medical history | Medical history structured by organ systems, medication, body weight, height, lifestyle (including smoking status, alcohol consumption, physical activity) |
| Physical status and<br>functional tests | Tinetti Mobility Test, Timed up & Go Test, Barthel Index (ADL), Lawton Instrumental Activities of Daily Living Scale (IADL), hand grip strength, anthropometric parameters, pulse wave velocity/arterial stiffness (Mobil-o-Graph), echocardiography, electrocardiography (ECG), spirometry, motion monitoring (Actigraph), dual-energy X-ray absorptiometry (DXA)<sup>1</sup>, |
| Psychological<br>screening tests | Mini Mental State Examination (MMSE), Digit Symbol Substitution Test (DSST)<sup>2</sup>, Center for Epidemiologic Studies Depression Scale (CESD) |
| Questionnaires | EPIC (food-frequency questionnaire), Gender Questionnaire, Pittsburgh Sleep Quality Index, Rapid Assessment of Physical Activity, SARC-F , SF-36 |
| Laboratory values<sup>3</sup> | Blood, serum or plasma: 25-hydroxyvitamin D, apolipoprotein A1, apolipoprotein B, basophiles, calcium, cortisol, creatinine, creatinine kinase, C-reactive protein, cystatin C, dehydroepiandrosterone, eosinophils, erythrocytes, ferritin, folic acid, gamma-glutamyltransferase, glucose 1, glucose 2<sup>4</sup>, glutamate, oxalacetate transaminase, glutamate-pyruvate transaminase, HbA1c, HDL-cholesterol, hematocrit, hemoglobin, homocysteine, international normalized ratio, iron, LDL- |
|  | cholesterol, leukocytes, lipoprotein (a), lymphocytes, magnesium, MCH, MCHC, MCV, monocytes, neutrophils, oestradiol, osteocalcin, partial thromboplastin time, RDW, sex hormone-binding globulin, testosterone, thrombocytes, thyroid-stimulating hormone, thyroxine, total-cholesterol, triglycerides, triiodothyronine, urea, uric acid, vitamin B12, zinc.<br>Urine: albumine, creatinine, desoxypyridinoline, teststrip: bilirubin, blood (erythrocytes), glucose, ketones, leukocytes, nitrite, pH value, protein, specific weight, urobilinogen<br>Dried blood cards: arsenic, brain derived neurotropic-factor, cadmium, chromium, fatty acids (C12:0, C14:0, C15:0, C16:0, C16:1n7, C17:0, C18:0, C18:1,t6-11, C18:1,c9, C18:1,c11, C18:2,n-6, C20:0, C18:3,n-6, C18:3,n-3, C20:1,n-9, C20:2,n-6, C22:0/C20:3,n-6, C20:4,n-6, C20:5,n-3, C24:0, C22:5,n-3, C22:6,n-3, unknown), HbA1c, hsCRP, lead, mercury, nickel, total-cholesterol |
| Genomics | Genome-wide single nucleotide polymorphism genotyping using the “Global Screening Array” (Illumina, Inc.); genome-wide DNA methylation profiling using the “Infinium MethylationEPIC” array (Illumina, Inc.) |
| Psychosocial questionnaire | Well-being, positive affect and negative affect, emotion regulation, stress, personality, control beliefs, domain-specific control, time perception, embitterment, loneliness, solitude, social activities, network structure, sexuality, risk behavior, etc. |
| Biobanking | Blood plasma & serum, urine, DNA extracted from EDTA-blood and buccal swaps |
| Cognitive tests | *Episodic memory* (Picture-Word-Task, Face-Profession-Task, Object Location Task, Scene-Encoding, Verbal learning and memory test), *Working memory* (Letter Updating, Spatial Updating, Number-N-Back), *Executive functioning / processing speed* (Multi-Source-Interference Task, Digit Symbol Substitutions Test<sup>2</sup>, Fluid intelligence (Letter series, Number series, Practical Problems), *Subjective Health Horizon Questionnaire* (SHH-Q) |
| Immunological assessment | Cryopreservation of whole blood (SmartTube system) or isolated PBMCs, and serum samples. Direct ex vivo staining of recent thymic emigrants (RTE, CD31+ CD45RA+ CD4+ T cells), TEMRA (CD45RA+ CD8+ T cells), Tregs (CD25bright CD127- CD4+ T cells), cytotoxic CD4+ T cells, amongst others using four different panels: 1) ImmunoCount Panel (CD45, CD3, CD56, CD19, CD16, CD14, CD123, CD1c); 2) RTE panel (CD3, CD4, CD8, CD45RA, CCR7, CD31, CD95, CD11a); 3) TREG panel (CD3, CD4, CD8, CD25, CD127); 4) Effector T cell panel (CD3, CD4, CD8, CD45RA, CCR7, SLAM-F7, IL-6R, CD57, PD-1). Panels were measured on MacsQuant 10 (Miltenyi), MacsQuant 16 (Miltenyi) or LSR II (BD) |

<sup>1</sup>only available for older group; <sup>2</sup>assessed at study visit 1 and 3, <sup>3</sup>blood samples were drawn after a fasting period of at least 8 hours (if not otherwise indicated), <sup>4</sup>post-load (75g glucose, 2h), not assessed in participants with known diabetes

With a focus on cardio-metabolic diseases in GendAge, we extended the broad range of data assessed in this area at baseline by echocardiography. Data on right and left ventricular and atrial morphology and systolic and diastolic function and vascular stiffness were obtained by a trained investigator (US) on a General Electric Vivid T8 R3 System and analyzed by a trained analyst (ET). 10% of echo analysis were controlled by an independent supervisor. The intraclass correlation coefficient (ICC) between analyst and independent supervisor was 0.87 (LV IVSd: 0.86, LVEDV Biplane: 0.95, LVEF Biplane: 0.87, E/A: 093). LVEF was higher in women than in men, in agreement with most recent publications, and probably partially due to higher rate of MI in men^34^.

Peripheral blood mononuclear cells were prepared from 903 participants at follow-up and frequencies as well as absolute counts of recent thymic emigrants (RTEs), T_EMRA_ effector T cell subsets (T_EMRA_) and cytotoxic CD4^+^ T cells were directly assessed. While RTEs are known to decrease with ageing^35^, alterations in T_EMRA_ and specialised cytotoxic CD4^+^ T cell compartments can be indicative of age-related perturbations of systemic T cell immunity^36^.

Similar to baseline, we determined numerous routine laboratory parameters from blood and urine (Table 2), and also stored blood plasma/serum and urine samples for future analyses.

Genomic DNA was already extracted from EDTA-blood and buccal swab samples from GendAge participants, which will be used e.g. for the profiling of genome-wide DNA methylation signatures and new genome-wide single nucleotide polymorphism genotyping experiments (Table 2). In-between the two assessment days at the Charité, participants were asked to fill out a comprehensive psychosocial take-home questionnaire and return this at their second Charité visit. Moreover, another wave of cognitive assessments (see Table 2) carried out by the Max Planck Institute for Human Development (MPIB) has been tightly linked to the GendAge assessment of BASE-II participants. The cognitive session (= third study visit) was followed 7 days after the second medical examination and lasted about 4.5 to 5 hours. Subjects were tested in groups of 4–6 individuals. The cognitive battery included 17 measures of learning and memory performance, attention/processing speed, working memory, executive functioning, and perceptual speed (see Table 2). Within the week between study visit 2 (Charité) and 3 (MPIB) accelerometers (ActiGraph wGT3X-BT) have been used to track participants’ physical activity and sleep in a subset of our participants (N = 750).

After the cognitive session, participants were invited to take part in a one-to-one interview on a different day. This additional individual assessment took up to 60 minutes and serves as a cohort comparison between the BASE and BASE-II study populations. This additional data collection will also contribute to the BASE-II cognitive waves, allowing us to further investigate individual differences in aging trajectories (for an overview, refer to^20^).

Furthermore, and as part of a collaboration with the Lifebrain study, a consortium of European studies funded by the EU Horizon 2020 Framework Programme^37^, we collected blood samples using dried blood cards, in order to determine laboratory parameters with identical methods used for all Lifebrain participating sites. Lifebrain aims at identifying determinants of healthy lifespan development by integrating and harmonizing data and results from 11 large and predominantly longitudinal European samples from 7 countries.

This has yielded a database of fine-grained measures focusing on brain and cognition from more than 7,000 individual participants.

The GendAge study was approved by the Ethics Committee of the Charité– Universitätsmedizin Berlin (approval number EA2/144/16) and all participants gave written informed consent. GendAge is registered in the German Clinical Trials Register (Study-ID: DRKS00016157). The cognitive battery was approved by the Ethics Committee of the Max-Planck-Institute and the genomics experiments were approved by the Ethics Committees of the Charité (approval number EA2/144/16) and the University of Lübeck (approval numbers AZ19–390A and 19–391A).

## What has been found?

GendAge was initiated in 2017 with a focus on sex and gender differences in the ongoing analyses of baseline BASE-II data in the area of cardio-metabolic risk factors and disease, some of which we would like to highlight here. The metabolic syndrome (MetS) combines the five cardiovascular risk factors of high waist circumference, low high density lipoprotein cholesterol, high fasting blood glucose, high triglycerides, and high systolic/diastolic blood pressure to diagnose the MetS based on cutoff values for each of the risk factors^38^. While this approach is certainly useful in the clinical setting, it does not consider the full range of information of the continuously measured risk factors. We therefore applied confirmatory factor analysis to establish a latent metabolic load factor (MetL), representing a single continuous measure based on the five MetS risk factors. The MetL was on average significantly higher in men compared to women, and the factor was associated with more morbidity and poorer kidney function in both men and women, and with lower performance on a measure of cognitive function in men^7^. Strong evidence suggests that higher levels of Lipoprotein (a) [Lp(a)] are an independent risk factor for coronary heart disease^39^. Interestingly, at the same time higher Lp(a) is associated with a reduced type 2 diabetes (T2D) risk, a finding which has been reported from numerous studies, including BASE-II^6,40,41^. We recently analyzed the relationship between Lp(a) and cognitive performance assessed with the *Consortium to Establish a Registry for Alzheimer’s Disease* (CERAD)-Plus test battery. The results suggested that lower Lp(a) levels associate with better cognitive performance, at least in men^8^. The analysis of arterial stiffness data from BASE-II assessed at baseline revealed sex differences in measures of arterial wave reflection, with an on average higher augmentation index in women than in men, a measure of endothelial function of the small and medium arteries. The results also suggested that women with exogenous intake of sex hormones for oral contraception and suppressed endogenous estradiol levels had higher levels of the augmentation index than non-users^42^. T2D is an age-associated disease which is reflected by the 4.8% increase in the proportion of affected BASE-II participants within the 7.35 (SD ± 1.46) years between baseline^6^ and follow-up assessments. This is even more dramatic since almost one third of the participants was not aware of the disease (Table 1). In addition being a risk factor for cardiovascular disease and cognitive decline, a recent analysis of BASE-II data suggested that the capacity to regulate the glucose metabolism is associated with well-being among older men^43^.

The immunological screening has so far revealed significantly higher frequencies of RTEs in women as compared to men, indicating a higher thymic T cell production even at the advanced ages of the GendAge participants. In men, more CD45RA^+^ re-expressing TEMRAs were detected than in women (Table 1). These cells are associated with chronic viral infections (e.g. CMV) and can serve as a signature of immune-senescence^44^. We found no significant difference in the frequencies of cytotoxic CD4^+^ T cells. Together, these preliminary findings confirm the better immune status of aged women as compared to men. A detailed analysis of the datasets will identify additional correlates of sex and gender, aging, and the immune system.

In GendAge, we have developed a retrospective gender score taking BASE-II baseline data reflecting sociocultural aspects (e.g., level of education, marital status, and chronic stress) into account. This gender score (GS-I) was associated with a number of clinical and psychosocial variables and performed better in predicting differences in a subset of variables compared to biological sex^22^. As mentioned above, we have implemented a comprehensive gender questionnaire as part of the follow-up assessments described here, including not only the variables required to re-calculate the GS-I based on the follow-up data, but also the items to develop an adapted version of the gender questionnaire proposed by Pelletier and colleagues^21^. This gender questionnaire will be central to develop a second gender score (GS-II) and to reach the GendAge goals.

## What are the main strengths and weaknesses?

The BASE-II follow-up assessments covered most of the medical, psycho-social and cognitive domains and variables assessed at baseline, and thereby taking the BASE-II characteristic of an exceptionally broad and in-depth data collection to a next, longitudinal level. In addition, and in the context of the GendAge focus on cardio-metabolic disease, we extended the assessments in this area e.g. by including high-quality echocardiography resulting in a unique data collection. This strength with respect to comprehensive and longitudinal data offers the potential to answer a number of questions that are of crucial relevance for the health of old women and men. Thus, GendAge will make important contributions for improvements in understanding the health and well-being of older adults in both genders. BASE-II was initiated as a multidisciplinary study with expertise in a broad range of fields relevant for aging research (e.g. internal medicine and geriatrics, biology, psychology, genetics, immunology, socio-economics, and now in GendAge further extended by socio-cultural aspects of gender). The past ten years of BASE-II research have shown that multidisciplinary collaboration is not only a statement of intent, but a fruit-bearing working posture and a clear strength of BASE-II.

Sampling bias is a challenge which cohort studies have to deal with, and this is especially an issue in the follow-up of older study populations such as the older group of BASE-II participants. To address this, we have made a considerable effort (e.g. offering a taxi service for participants not able to travel independently) to include as many participants in the followup as possible. Additionally, and similar to baseline, we are able to systematically quantify the sampling bias and even account for it when it comes to the question of generalizability of study results to a population as a whole (e.g., Berlin or Germany), due to the evaluation of selectivity and representativeness via the German Socio-Economic Panel Study (SOEP)^1^. Despite these possibilities we cannot rule out the possibility of a selection bias completely, which certainly is a weakness of this study, a weakness that applies to all cohort studies relying on voluntary participants who have been non-randomly recruited. With our direct comparability to the national representative SOEP study, we are in a position though to quantify the amount of selectivity and, if need, take measures to correct and adjust our results.

## Can I get hold of the data? Where can I find out more?

More details on GendAge can be found at https://gendage.charite.de/en/ and information on the BASE-II as a whole is available at https://www.base2.mpg.de/en. BASE-II has a tradition of sharing data and biobank samples in joint collaborative projects which will be continued with respect to BASE-II data assessed in GendAge. Interested groups are invited to contact the study coordinating PI Ilja Demuth at for the data-sharing application form. Each application will be reviewed by the GendAge PIs (currently ID, VRZ and DG) and the decision communicated to the applicants usually within 4 weeks of submission.

## Acknowledgements

We are deeply indebted to all individuals of the BASE-II cohort who have agreed to participate in the follow-up assessments. We would further like to thank all staff members and students involved in the medical examinations of participants and documentation, especially Nora Berger, Janina Dombrowski, Mergim Domuzeti, Elisa Dreißig, Ilona Enarovic, Anthony Ganswindt, Thomas Grenkowitz, Anna Hunold, Ilias Katsianas, Yoo-Ri Kim, Jörn Kiselev, Paula Krull, Christine Kytmannow, Elisa Lemke, Julia Mätzkow, Charlotte Mentzel, Nadja Mielke, Narantuya Mishig, Angela Motz, Eduard Nitschke, Aabi Okute, Danai Pantelakis, Sophie Poser, Johanne Spieker and Taleka Vollmar (all Charité Universitaetsmedizin Berlin, Biology of Aging group); and Luisa Lüth, Mariebelle Kaus, Isabel Ganter, Marlene Rosendahl, Jasmin Boneberger, Antonia Sprenger, Hania El-Kersh (all Humbold University, Department of Psychology), and Kirsten Becker for the excellent daily organization of the running study within and between the research units, the Telefonstudio for their daily contact with the participants, Martin Becker for the continuous quality checks of our sample and Berndt Wischnewski for updating the database and cognitive battery (all Max Planck Institute for Human Development).

We kindly acknowledge the excellent cooperation with the Central Biomaterial Bank, the joint core facility of the Charité-Universitätsmedizin Berlin and the Berlin Institute for Health (BIH). Furthermore, we appreciate the great support by the BIH-REDCap team and especially from Andreas Hetey.

All co-workers listed above have confirmed their agreement to be acknowledged in this paper.

## Additional BASE-II/GendAge investigators

Nikolaus Buchmann^i^, Peter Eibich^ii^, Friederike Kendel^iii^, Maximilian König^iv^, Christina M. Lill^v,vi^, Maike Mangold^vii^, Ahmad Tauseef Nauman^iii^, Kristina Norman^viii,ix^, Graham Pawelec^x,xi^, Elisabeth Steinhagen-Thiessen^iv^, Sarah Toepfer^iv^, Valentin Max Vetter^iv^, Gert G. Wagner^xii^, Ursula Wilkenshoff^i^, Kilian Wistuba-Hamprecht^xiii^

^i^Charité; Department of Cardiology, Charité - University Medicine Berlin (Campus Benjamin Franklin), Berlin, Germany; ^ii^Max Planck Institute for Demographic Research, Rostock, Germany; ^iii^Berlin Institute for Gender in Medicine, Charité - Universitätsmedizin Berlin; ^iv^Charité - Universitätsmedizin Berlin, corporate member of Freie Universität Berlin, Humboldt-Universität zu Berlin, and Berlin Institute of Health; Department of Endocrinology and Metabolism, Berlin, Germany; ^v^Section for Translational Surgical Oncology and Biobanking, Department of Surgery, University of Lübeck and University Medical Center Schleswig-Holstein, Campus Lübeck, 23552 Lübeck, Germany; ^vi^Ageing Epidemiology Research Unit, School of Public Health, Imperial College, London SW71, UK; ^vii^Regenerative Immunology and Aging, BIH Center for Regenerative Therapies, Charité Universitatsmedizin Berlin, Berlin, Germany; ^viii^German Institute of Human Nutrition, Department of Nutrition and Gerontology, Potsdam-Rehbruecke (DIfE), Germany; ^ix^Charité - Universitätsmedizin Berlin, Forschungsgruppe Geriatrie am EGZB, Berlin, Berlin, Germany; ^x^Department of Immunology, University of Tübingen, Tübingen, Germany; ^xi^Health Sciences North Research Institute, Sudbury, ON, Canada; ^xii^German Socio-Economic Panel Study (SOEP); ^xiii^Division of Dermatooncology Department of Dermatology, University of Tübingen, Tübingen, Germany.

